# Dissection of the ‘Virgin-soil’ epizootic of African swine fever in Mizoram, a Northeast state of India

**DOI:** 10.1101/2024.09.09.24313373

**Authors:** Santhalembi Chingtham, Freda Lalrohlui, Abigail Remlalfakawmi, C. Neihthangpuii, Esther Lalzoliani, Parimal Roychoudhury, Prashant Kumar Subudhi, Tapan Kumar Dutta

## Abstract

This article aims to analyse the first-time emergence of African swine fever (ASF) in Mizoram. We collated the outbreak data and identified the time and locations of ASF emergence. To understand the impact of the outbreak, we have calculated the morbidity rate, mortality rate, case fatality rate (CFR) and overall pig depopulation rate. We identified that ASF emerged in total 178 locations in all 11 districts of Mizoram between March-July 2021, after a peak in June, and the disease continued to spread till the end of 2021 before re-emergence in March 2022. The overall morbidity rate and mortality rate of ASF between March-July 2021 in Mizoram were estimated to be 33.8% and 31.1%, while the average morbidity rate and the mortality rate of 10 districts were found to be 29.5% and 27.6% respectively. Overall CFR of ASF between March-July 2021 was estimated to be 92.1% and the average of all 11 districts was found to be 96.1%. Toward the end of 2021, the mortality rate increased by a total of 42.7% change and an average of 25.3% change. We estimated an overall 70% depopulation of susceptible pigs by disease and culling by end of 2021. This report is evidence that ASF has remained catastrophic and, keeping in view the complex nature and history of the virus on genotype adaptability associated with a high propensity for transboundary expansion, the virus continues to be a threat.

## Introduction

African swine fever (ASF) is a disease of domestic, feral and wild *Sus scrofa* (pig) notifiable to World organisation for Animal Health (WOAH) [1], bearing serious negative implications on national and international trade and economy [2]. It is caused by infection with a complex enveloped DNA virus called the African swine fever virus (ASFV) - the only known DNA arbovirus and the only member of the genus *Asfivirus,* family *Asfaviridae* [1,3,4]. First detected in Kenya in 1909 [5], ASF is endemic to the Sub-Saharan Africa [12] and the virus is known to be maintained in the eastern and southern Africa in a natural sylvatic cycle of vertebrate reservoir hosts - the African wild suids (Warthogs, bushpigs, red river hog and giant forest hogs), and arthropod hosts - argasid ticks of *Ornithodorous moubata* complex that act as biological vector [1,3,7]. The original ASF virus ecology is complex in the sense that the infection pattern of the wild suid population and soft ticks varies with geographic location and virus genotype, and sporadic domestic cycle runs independently from and parallel to the sylvatic cycle with wild infected ticks more likely serving as accidental bridge [3]. Trans- and inter-continental expansion of the domestic cycle that began with ASF appearance in Portugal in 1957 and subsequently in other parts of Iberia and South America, Caucasus and Russia, to the rapid expansion in Eurasia recently is largely attributed to the horizontal transmission of the virus between pigs in domestic and wild population of *Sus scrofa* instigated by global trade and other cross-continental human activities, thereby, endangering the wild population of *Sus scrofa* while also magnifying the risk of virus maintenance in the wild population of susceptible suids and ticks in non-endemic regions [3,7,8]. The role of infected wild *Sus scrofa* (wild boar) in the ASF epidemiology is evident in the recent ASF outbreaks in Europe and parts of Asia where the transmission of ASF is reported to be dependent largely on wild boar population coupled with a low biosecurity in pig production systems [8]. These reports strongly imply that the complex yet stable ASF virus ecology and ASF epidemiology culminates to a challenge on risk and impact mitigation of ASF once the virus is introduced and established in a region. According to the biweekly update of WOAH report on global ASF situation 74 countries have been affected during 2005 and 2022, and the disease has been reported as present in 45 countries affecting 1,191,000 pigs and more than 37,000 wild boars for the period between January 2020 and December 2022 [9]. In Asia, ASF emerged for the first time in China in August 2018 and by 2021, 16 countries were already affected including India [8, 10].

Infection with ASF virus is manifested in multi syndromic forms of varying degrees of severity from hyperacute to chronic and asymptomatic depending on the virus strains, geographic location of incidence, hosts, vectors and other factors [3,7,4,11]. Incidence, particularly for the first time in a new location, is commonly associated with fatal outcomes of the most severe form in domestic, feral or wild *Sus scrofa* manifesting up to 100% mortality and case fatality rates [3]. Symptoms and lesions in the course of the disease, if not preceded by death, commonly involve haemorraghic manifestations that have often led to confounding judgements in a first-time occurrence due to the resemblance to other endemic porcine haemorrhaghic diseases like the classical swine fever, thereby impeding the process of specific impact mitigation of ASF [3,4].

In India, first laboratory confirmed ASF in domestic pigs was reported in January 2020 in 2 districts of Arunachal Pradesh and 5 districts of Assam [10, 12], which are two neighboring states in the northeast region of the country. Arunachal Pradesh also shares national border with Nagaland, and international border with China, Myanmar and Bhutan, while Assam shares international border with Bangladesh and Bhutan, and national border with 5 other states in the northeast including Mizoram. Post the outbreaks in the two Northeastern states, multiple new outbreaks were reported in 5 different states within the Northeast region - Meghalaya and Manipur between August 2020 to November 2020, Nagaland and Mizoram in March 2021, and Sikkim in early March 2022 [12].

Mizoram is a state in the northeast region of India covering an area of approximately 21,087 sq km of mostly hilly terrain with 89% forest coverage [13], shares 3 national borders with Assam (123 km), Tripura (277 kms) and Manipur (95 kms) [14]. The state also shares 404 km and 318 km long international borders with Myanmar and Bangladesh respectively. The state is an agrarian economy with backyard piggery being one of major sources of local livelihood [15]. According to the 20th Livestock Census released in 2019 by the Department of Animal Husbandry and Dairying, Government of India, the total population of pigs in Mizoram was 2,83,021 (shown in Table 1) which is 82.2% of the total livestock population in Mizoram in 2019 [16]. Mizoram has 11 districts - Aizawl, Lunglei, Champhai, Mamit, Saitual, Khawzawl, Hnahthial, Lawngtlai, Serchhip, Kolasib, and Siaha [17]. In this article, we aim to provide an account on the ASF outbreak and attempt to analyze the impact of the first-time emergence of ASF in Mizoram while also highlighting the consolidation of risk factors, and the future threats. There is no historical record on prevalence of ASFV infection prior to the outbreak described in this article, and although a study on genetic characterization of ASFV from the same outbreak has been reported earlier [18], this is the first description of ASF in Mizoram. In this article, ASF outbreak events in 2021 have been collectively referred to as ‘first outbreak ‘and the events in 2022 as ‘second outbreak ‘and the focus of analysis and discussion is kept on the events in 2021.

**Table 1.**
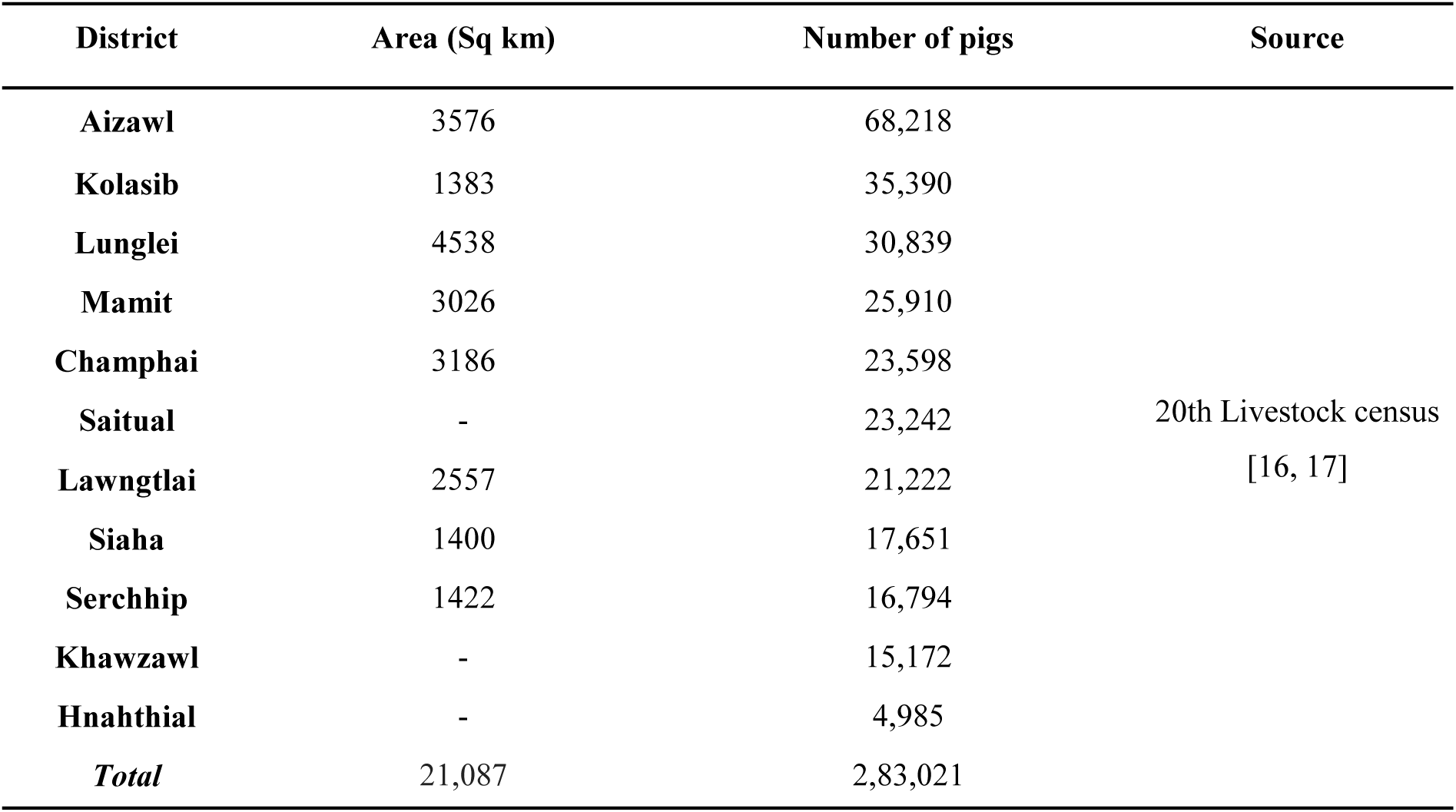
Mizoram Pig population in 2019.

## Methodology

Data on the ASF outbreaks within Mizoram state was acquired from the State Department of Veterinary and Animal Husbandry, Government of Mizoram. The data was curated, and it consisted of the number of susceptible animal at specified epicentre locations within the districts of Mizoram state, number of suspected and/or laboratory confirmed cases, number of ASF induced deaths, number of culled pigs and number of families affected. Infection status or number of cases reported was based on clinical manifestation and laboratory confirmation of representative specimens. Initial laboratory confirmation by real-time PCR was performed at the ICAR-National Institute of High Security Animal Diseases, Bhopal (Madhya Pradesh), a national reference laboratory, and Northeast Regional Disease Diagnosis Laboratory, Guwahati (Assam), and subsequent laboratory detection of new appearances at different locations were done by PCR and rapid antigen tests at the diagnostic facility of the department of Veterinary Microbiology, College of Veterinary Science and Animal Husbandry, CAU, Selesih, Aizawl, Mizoram.

### ASF epicentres and timeline of ASF appearance

From the available data, the number of locations with first notification of the appearance of ASF between March 2021 and June 2021 in each of the 11 districts of Mizoram were identified. The locations within each district were then grouped month-wise to understand the month-wise distribution and first appearance of ASF for each district. The month in which the ASF appearances peaked was also identified. To determine the first onset of ASF within each district, a date-wise timeline is created consisting of reported dates of onset in different locations of each district. A number of locations and dates with overlapping ASF onset were also identified.

### Estimation of morbidity rate, mortality rate, case fatality rate and culling rate

The ASF morbidity rate for each district was estimated using the available data on the number of susceptible pigs recorded for each affected location and the reported ASF cases during the period from first onset in March 2021 to July 2021. For the overall morbidity rate for the defined period in the whole of Mizoram the average of the estimates of each district is calculated. The total morbidity rate is calculated from the cumulative values of all districts. Except Saiha district, all 10 districts of Mizoram were considered for estimation of ASF morbidity rate. The formula below was used to calculate the morbidity rate:

*Morbidity rate (%) = {(Number of ASF cases)/ (Number of susceptible pigs)} x 100*

The ASF case fatality rate (CFR) for each district was estimated for the first half of the first outbreak using the available data on the number of reported ASF cases during the period from first onset in March 2021 to July 2021. For overall CFR (%) for the defined period in the whole of Mizoram the average of the estimates of each district is calculated. The total CFR (%) is calculated from the cumulative values of all districts. All 11 districts were included in the assessment of CFR. The formula below was used to calculate the morbidity rate:

*CFR (%) = {(Number of deaths due to ASF)/ (Number of ASF cases)} x 100*

ASF mortality rate for each district was estimated in percentage for two time periods in the first outbreak using the available data on the number of susceptible pigs recorded for each affected locations and the reported ASF mortality during the period from first onset in March 2021 to July 2021, and the period from first onset in March 2021 to December 2021. For overall mortality rate for the defined periods in the whole of Mizoram the average of the estimates of each district was calculated. The total mortality rate was calculated from the cumulative values of all districts. Except Saiha district, all 10 districts of Mizoram were considered for estimation of ASF mortality rate. The formula below was used to calculate the morbidity rate:

*Mortality rate (%) = {(Number of deaths due to ASF)/ (Number of susceptible pigs)} x 100*

Overall ASF induced age-wise mortality is also estimated for all 11 districts of both first outbreak in 2021 and second outbreak in 2022. The mortality was grouped under three age groups of below 3 months, between 3-8 months and above 8 months. The mortality rate in each age group was not determined because of unavailable data on the age-wise number of susceptible pigs.

### Estimation of culling rate and overall depopulation rate

Culling rate was estimated for each district for two time periods - March 2021 to July 2021 and March 2021 to December 2021 using the formula below. Except Saiha district, all 10 districts of Mizoram were considered for estimation of culling rate. The number of culled pigs were also categorised into five groups according to body weight for each district.

### Culling rate (%) = {(Number of culled pigs)/ (Number of susceptible pigs - Number of deaths due to ASF)} x 100

Overall depopulation status was estimated for each district for two time periods in the first outbreak - March 2021 to July 2021 and March 2021 to December 2021 using the data on number of deaths due to ASF infection and number of pigs culled.The depopulation rate was calculated using the estimated depopulation value and the number of susceptible pigs. The number and proportion of live pigs toward the end of 2021 were also estimated.

*Depopulation rate (%) = {(Number of deaths due to ASF+Number of culled pigs)/ (Number of susceptible pigs)} x 100*

### ASF mortality and families affected in first outbreak and second outbreak

To compare the district wise number of ASF induced pig mortality and families affected in the first and second outbreaks the percent proportion of mortality and affected families were calculated for each district for the first outbreak in 2021 and second outbreak in 2022.

## Results

### ASF epicentres and timeline of ASF appearance

ASF appeared in a total of 178 different locations for the first time in all 11 districts of Mizoram with different and overlapping dates of onset during March 2021 to July 2021. ASF appeared in 78 locations in Aizawl district, 5 in Champhai, 6 in Hnahthial, 4 in Khawzawl, 3 in Kolasib, 3 in Lawngtlai, 17 in Lunglei, 24 in Mamit, 12 in Saitual, 25 in Serchhip, and 1 in Siaha. The onset of ASF outbreak was seen in 6 locations within 6 districts in the month of March, 25 locations within 7 districts in April, 36 locations in 6 districts in May, 74 locations in 7 districts in June, and 37 locations in 10 districts in July. The breakdown of month-wise appearances of ASF for each district is presented in Table 2. Total ASF appearances peaked in the month of June 2021 (Figure 1). A total of 87 dates from 9th March 2021 to 31st July 2021 have been recorded with the onset of ASF in the above locations. Total 130 locations of 10 districts (except Siaha) showed onset of ASF on 33 overlapping dates - 0 overlapping onset date in the month of March 5 in April with 13 locations within 5 districts, 4 in May with 24 locations within 6 districts, 16 in June with 67 locations within 7 districts, and 8 in July with 26 locations within 10 districts (Figure 1). The locations of the first-time onset of ASF in every district are shown in figure 1.

**Figure 1:**
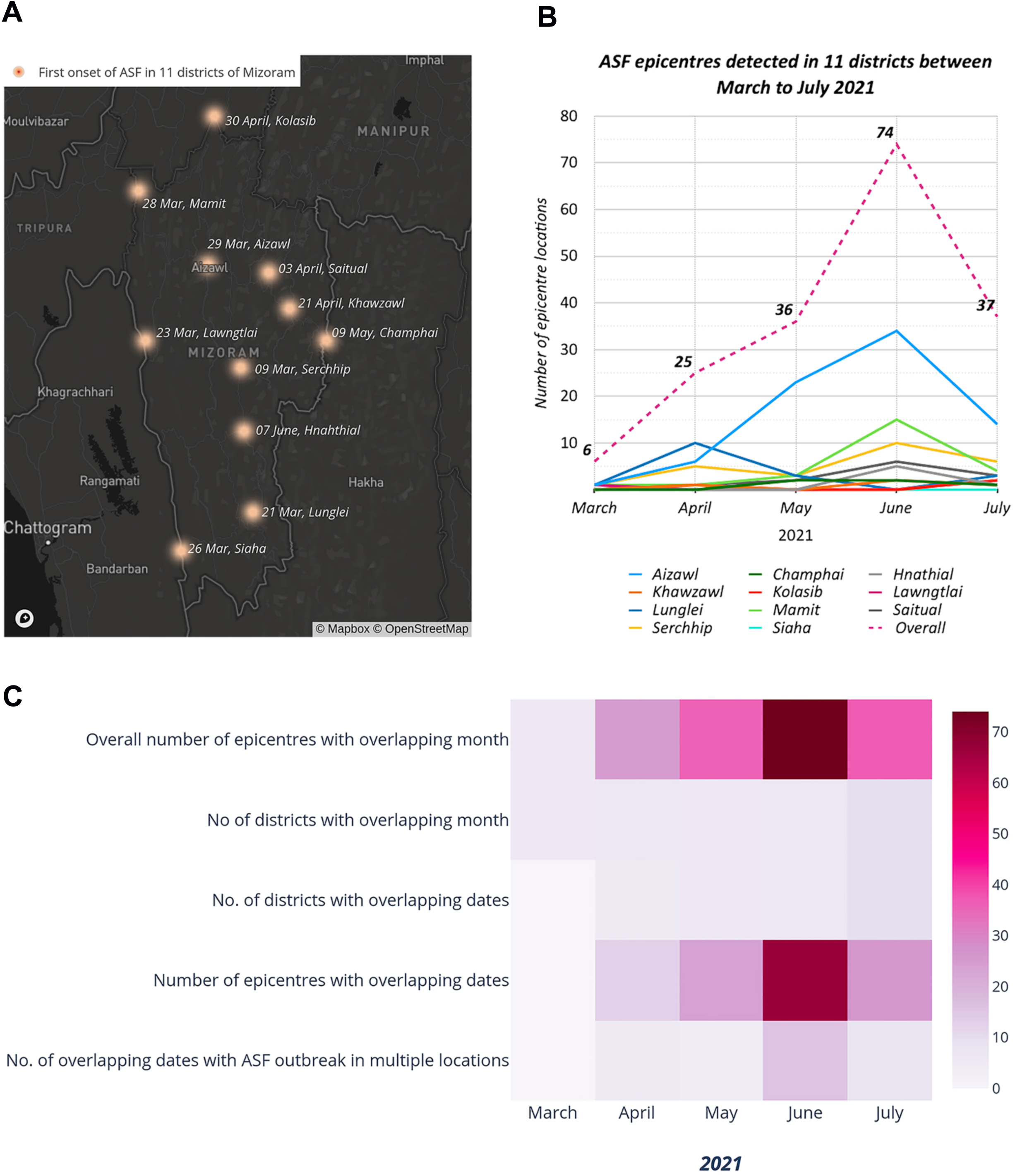
ASF epicentres and timeline of ASF appearance in Mizoram. A- Location of first onset of ASF recorded in 11 districts, B. Month-wise emergence of ASF in 11 districts, C- Heatmap showing overlapping outbreaks.

**Table 2.** ASF epicentres recorded in 11 districts between March and July 2021.

| **District** | **Aizawl** | **Champhai** | **Hnathial** | **Khawzawl** | **Kolasib** | **Lawngtlai** | **Lunglei** | **Mamit** | **Saitual** | **Serchhip** | **Siaha** | **Overall number of**<br>**epicentres with**<br>**overlapping month** | **No of districts**<br>**with overlapping**<br>**month** |
| --- | --- | --- | --- | --- | --- | --- | --- | --- | --- | --- | --- | --- | --- |
| **March** | 1 | 0 | 0 | 0 | 0 | 1 | 1 | 1 | 0 | 1 | 1 | 6 | 6 |
| **April** | 6 | 0 | 0 | 1 | 1 | 0 | 10 | 1 | 1 | 5 | 0 | 25 | 7 |
| **May** | 23 | 2 | 0 | 0 | 0 | 0 | 3 | 3 | 2 | 3 | 0 | 36 | 6 |
| **June** | 34 | 2 | 5 | 2 | 0 | 0 | 0 | 15 | 6 | 10 | 0 | 74 | 7 |
| **July** | 14 | 1 | 1 | 1 | 2 | 2 | 3 | 4 | 3 | 6 | 0 | 37 | 10 |
| **Total** | 78 | 5 | 6 | 4 | 3 | 3 | 17 | 24 | 12 | 25 | 1 | 178 |  |

### Estimation of morbidity rate, mortality rate, and case fatality rate

The overall morbidity rate of ASF in Mizoram in the initial period of the first outbreak (March-July 2021) was estimated to be 33.8% and the average morbidity rate of 10 districts was found to be 29.5%. The overall mortality rate was determined to be 31.1% while the average mortality rate of 10 districts was found to be 27.6%, the highest was in Lunglei with 41.8% while the lowest was in Kolasib with 2.13%. ASF mortality rate was also evaluated for the whole of first outbreak period (March-Dec 2021) and total mortality rate was found to be 52.63% while the average mortality rate of 10 districts was 48.2%, the highest was in Lawngtlai with 81.6% and the lowest was from Kolasib with 13.8%. The mortality rate increased by a total of 42.7% change and an average of 25.3% change toward the end of 2021 (Figure 2), highest rise was observed in Lawngtlai with a 68% change while lowest rise was observed in Mamit with an 8.9% change in mortality rate from mid-2021 to end of 2021. Siaha district reported 4 cases and 4 deaths during March-July 2021 and 313 deaths by the end of 2021, however, the district could not be included in the assessment of morbidity rate and mortality rate as the data on number of susceptible pigs was not available for the district. Overall case fatality rate (CFR) of ASF in Mizoram in the initial period of the first outbreak (March-July 2021) was estimated to be 92.1% and the average CFR of all 11 district was found to be 96.1%, with highest CFR of 100% observed in 6 districts - Hnahthial, Khawzawl, Saitual, Kolasib and Siaha while a lowest of 76.5% seen in Mamit district. The district-wise breakdown of morbidity, mortality, morbidity rate, mortality rate and CFR are summarized in Table 3. Due to lack of information on the true number of cases for the period during March-December 2021, the CFR could not be estimated for this period. The morbidity rate, mortality rate and CFR of all assessed districts and their average are shown in figure 2. Of the total ASF induced mortality of n=33,417, 30.2% (n=10,093) was comprised of pigs in the age group below 3 months, 34.6% (n=11,552) in the age group between 3-8 months and 35.2% (n=11,772) in the age group above 8 months of all 11 districts (Figure 2). Age-wise mortality rate could not be calculated because of the missing data on age-wise number of susceptible pigs.

**Figure 2:**
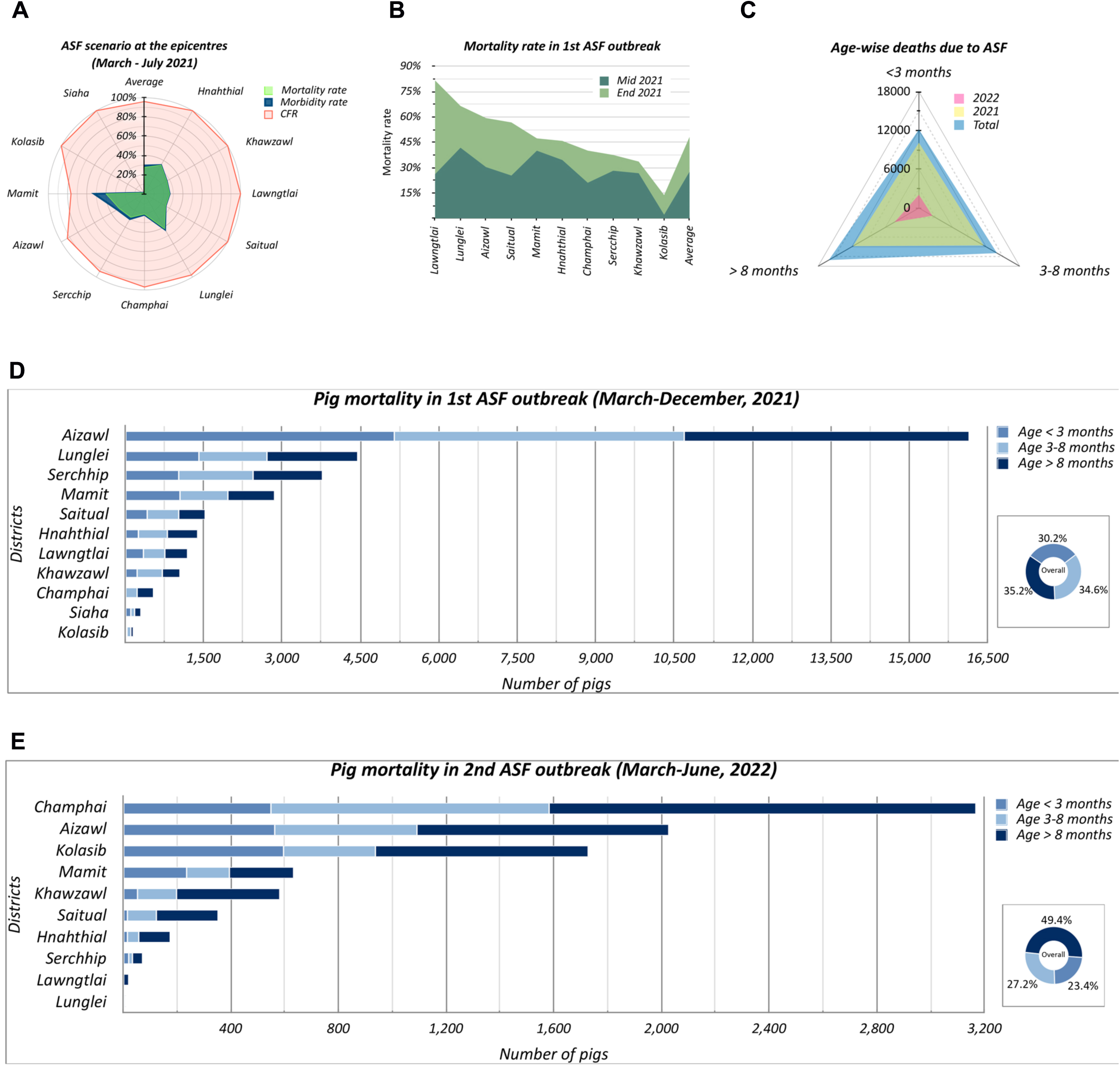
Impact of ASF outbreak in Mizoram. A- Morbidity rate, mortality rate and case fatality rate estimated for the outbreak during March 2021 to July 2021, B- District-wise percent change of ASF mortality rate from mid-2021 to end 2021, C- Overall age-wise, mortality observed in 2021 and 2022, D- District-wise and age-wise ASF-induced mortality in 2021, E-District-wise and age-wise ASF-induced mortality in 2022.

**Table 3:** ASF scenario in 2021 outbreak.

| District | No. of susceptible pigs | March to July 2021 |  |  |  |  | March to Dec 2021 |  |
| --- | --- | --- | --- | --- | --- | --- | --- | --- |
|  |  | Clinically determined cases (Morbidity) | Morbidity rate | Deaths due to infection (mortality) | Mortality rate | CFR | Deaths due to infection (mortality) | Mortality rate |
| Aizawl | 27,197 | 8,907 | 32.7% | 8,253 | 30.3% | 92.7% | 16,143 | 59.4% |
| Sercchip | 10,082 | 3,049 | 30.2% | 2,841 | 28.2% | 93.2% | 3,782 | 37.5% |
| Lunglei | 6,681 | 2,863 | 42.9% | 2,793 | 41.8% | 97.6% | 4,443 | 66.5% |
| Mamit | 6,022 | 3,153 | 52.4% | 2,411 | 40.0% | 76.5% | 2,850 | 47.3% |
| Khawzawl | 3,118 | 833 | 26.7% | 833 | 26.7% | 100.0% | 1,048 | 33.6% |
| Hnahthial | 3,043 | 1,051 | 34.5% | 1,051 | 34.5% | 100.0% | 1,395 | 45.8% |
| Saitual | 2,708 | 686 | 25.3% | 686 | 25.3% | 100.0% | 1,537 | 56.8% |
| Lawngtlai | 1,458 | 381 | 26.1% | 381 | 26.1% | 100.0% | 1,190 | 81.6% |
| Champhai | 1,367 | 297 | 21.7% | 288 | 21.1% | 97.0% | 548 | 40.1% |
| Kolasib | 1,219 | 26 | 2.1% | 26 | 2.1% | 100.0% | 168 | 13.8% |
| Siaha | - | 4 | - | 4 | - | 100.0% | 313 | - |
| *Total* | 62,895 | 21,250 |  | 19,567 |  |  | 33,417 |  |
| *Average* |  |  | 29.5% |  | 27.6% | 96.1% |  | 48.2% |

In the second outbreak between March - June 2022 total mortality of 8770 (ASF induced deaths) was observed, however the morbidity rate, mortality rate, and CFR of ASF in the second outbreak could not be calculated because of unavailable data on the number of susceptible pigs recorded for each affected locations and the number of active cases. Out of total ASF induced mortality of 8770, 23.4% (n=2,048) was comprised of pigs in the age group below 3 months, 27.2% (n= 2,388) in the age group between 3-8 months, and 35.2% (n=4,334) in the age group above 8 months of all 11 districts (Figure 2). Age-wise mortality rate could not be calculated because of the missing data on age-wise number of susceptible pigs.

### Estimation of culling rate and overall depopulation rate

A total of 6,360 pigs were culled during the period from March to July 2021 and a total of 11,177 pigs were culled in the period from March to December 2021. Overall culling rates of 10 districts in both the time periods were estimated to be 14.7% and 37.1% respectively, while the average culling rates of 10 districts in both the time periods were estimated to be 11.7% and 24.5% respectively. Four districts - Lawngtlai, Champhai, Khawzawl, Kolasib had 0% culling rate at all the locations of ASF outbreaks in 2021 and Mamit district showed the highest culling rate of 88.5% end of 2021. A total of 44,594 pigs were depopulated (disease+culling) in Mizoram during the first outbreak of ASF (Table 4). Overall depopulation rate in Mizoram was estimated to be 41.2% in the period from March to July 2021 and 70.2% in the period from March to December 2021, while the average depopulation rate of 10 districts in both the time periods were estimated to be 35.4% and 60.1% respectively. A total of 441 pigs were depopulated from Siaha district, however the district was not included in the assessment of the district-wise and overall culling rate and depopulation rate because of unavailable data on the number susceptible pigs at the locations. District-wise and overall depopulation status of the first ASF outbreak are shown in figure 3. A total of 19,009 pigs (30.2%) were estimated to be alive by the end of 2021 out of 62,895 susceptible pigs. The district-wise status of live pigs by end of first outbreak is shown in Figure 3.

**Figure 3:**
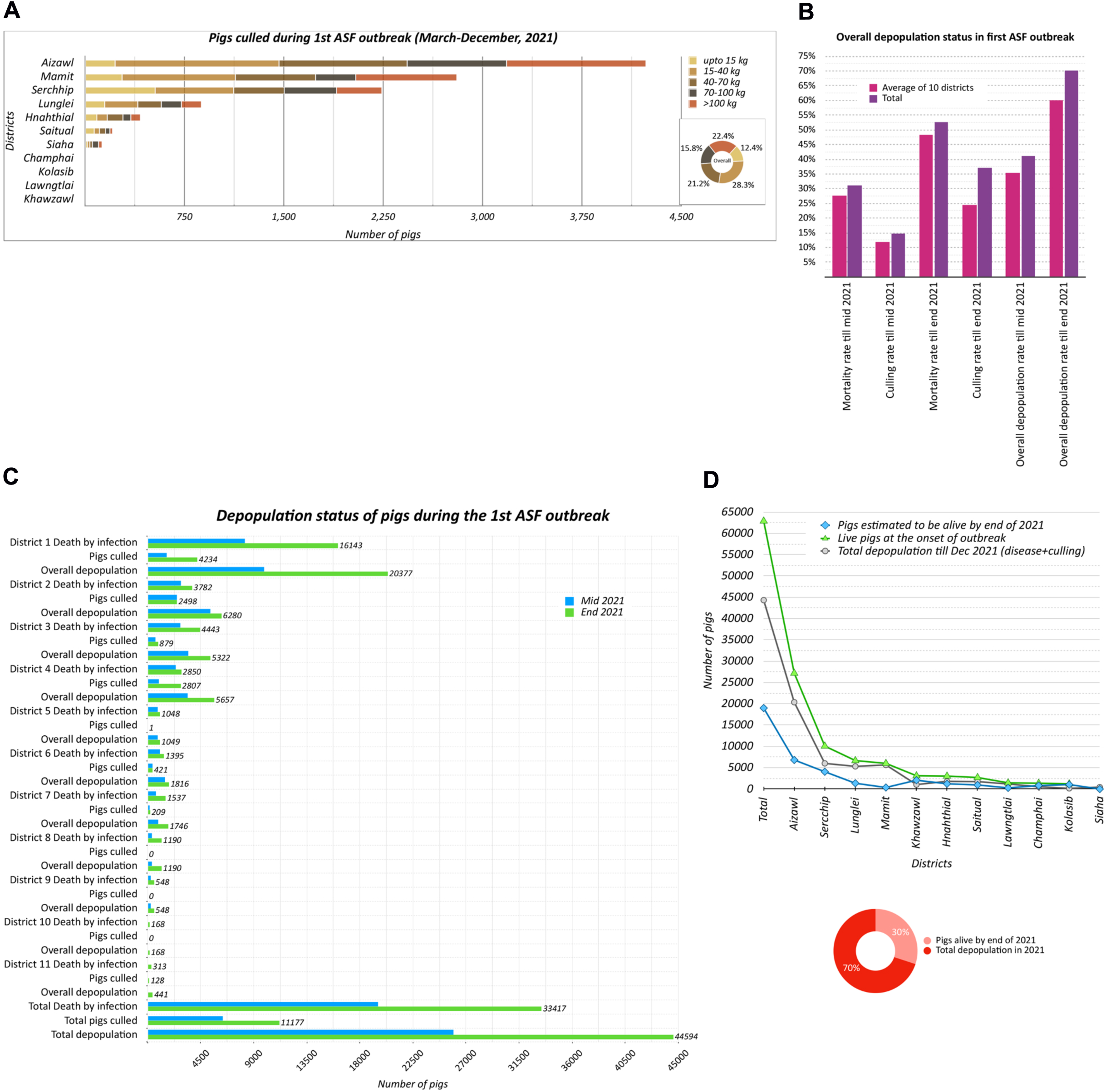
Depopulation status during the outbreaks of ASF in Mizoram in 2021. A- District-wise culling of susceptible pigs from March 2021 to December 2021, B- Overall culling rate and depopulation rate in Mizoram, C- District-wise number of ASF-induced deaths, culled pigs and overall number of pigs depopulated, D- Status of pig population in the affected locations in every district after depopulation.

**Table 4:** Pig depopulation status in 2021 outbreak of ASF.

| **District** | **Depopulation** | **Mid 2021** | **End 2021** |
| --- | --- | --- | --- |
| **District 1 (Aizawl)** | Death by infection | 8,253 | 16,143 |
|  | Pigs culled | 1,619 | 4,234 |
|  | Overall depopulation | 9,872 | 20,377 |
| **District 2 (Serrchip)** | Death by infection | 2,841 | 3,782 |
|  | Pigs culled | 2,498 | 2,498 |
|  | Overall depopulation | 5,339 | 6,280 |
| **District 3 (Lunglei)** | Death by infection | 2,793 | 4,443 |
|  | Pigs culled | 654 | 879 |
|  | Overall depopulation | 3,447 | 5,322 |
| **District 4 (Mamit)** | Death by infection | 2,411 | 2,850 |
|  | Pigs culled | 969 | 2,807 |
|  | Overall depopulation | 3,380 | 5,657 |
| **District 5 (Khawzawl)** | Death by infection | 833 | 1,048 |
|  | Pigs culled | 1 | 1 |
|  | Overall depopulation | 834 | 1,049 |
| **District 6 (Hnaththial)** | Death by infection | 1,051 | 1,395 |
|  | Pigs culled | 421 | 421 |
|  | Overall depopulation | 1,472 | 1,816 |
| **District 7 (Saitual)** | Death by infection | 686 | 1,537 |
|  | Pigs culled | 194 | 209 |
|  | Overall depopulation | 880 | 1,746 |
| **District 8 (Lawngtlai)** | Death by infection | 381 | 1,190 |
|  | Pigs culled | 0 | 0 |
|  | Overall depopulation | 381 | 1,190 |
| **District 9 (Champhai)** | Death by infection | 288 | 548 |
|  | Pigs culled | 0 | 0 |
|  | Overall depopulation | 288 | 548 |
| **District 10 (Kolasib)** | Death by infection | 26 | 168 |
|  | Pigs culled | 0 | 0 |
|  | Overall depopulation | 26 | 168 |
| **District 11 (Siaha)** | Death by infection | 4 | 313 |
|  | Pigs culled | 4 | 128 |
|  | Overall depopulation | 8 | 441 |
| **Total** | Death by infection | 19,567 | 33,417 |
|  | Total pigs culled | 6,360 | 11,177 |
|  | Total depopulation | 25,927 | 44,594 |

### ASF mortality and families affected in first outbreak and second outbreak

In the first outbreak between March-December 2021 48.3% of the total mortality (n=33,417) in Mizoram was from Aizawl district, 11.3% from Serchhip, 13.3% from Lunglei, 8.5% from Mamit, 3.1% from Khawzawl, 4.2% from Hnathial, 4.6% from Saitual, 3.6% Lawngtlai, 1.6% from Champhai, 0.5% from Kolasib, and 0.9% from Siaha. While in the second outbreak between March to June 2022., 23.1% of the total mortality (n=8,770) in Mizoram was from Aizawl district, 0.8% from Serchhip, 0.1% Lunglei, 7.2% from Mamit, 6.6% from Khawzawl, 2.0% from Hnahthial, 4.0% from Saitual, 0.2% from Lawngtlai, 36.1% from Champhai and 19.7% from Kolasib. The total ASF induced mortality from both the outbreaks is 42,187, out of which 46% was from the period of March to July 2021, 33% from August to December 2021 and 21% from March to June 2022. These findings are presented in figure 4.

**Figure 4:**
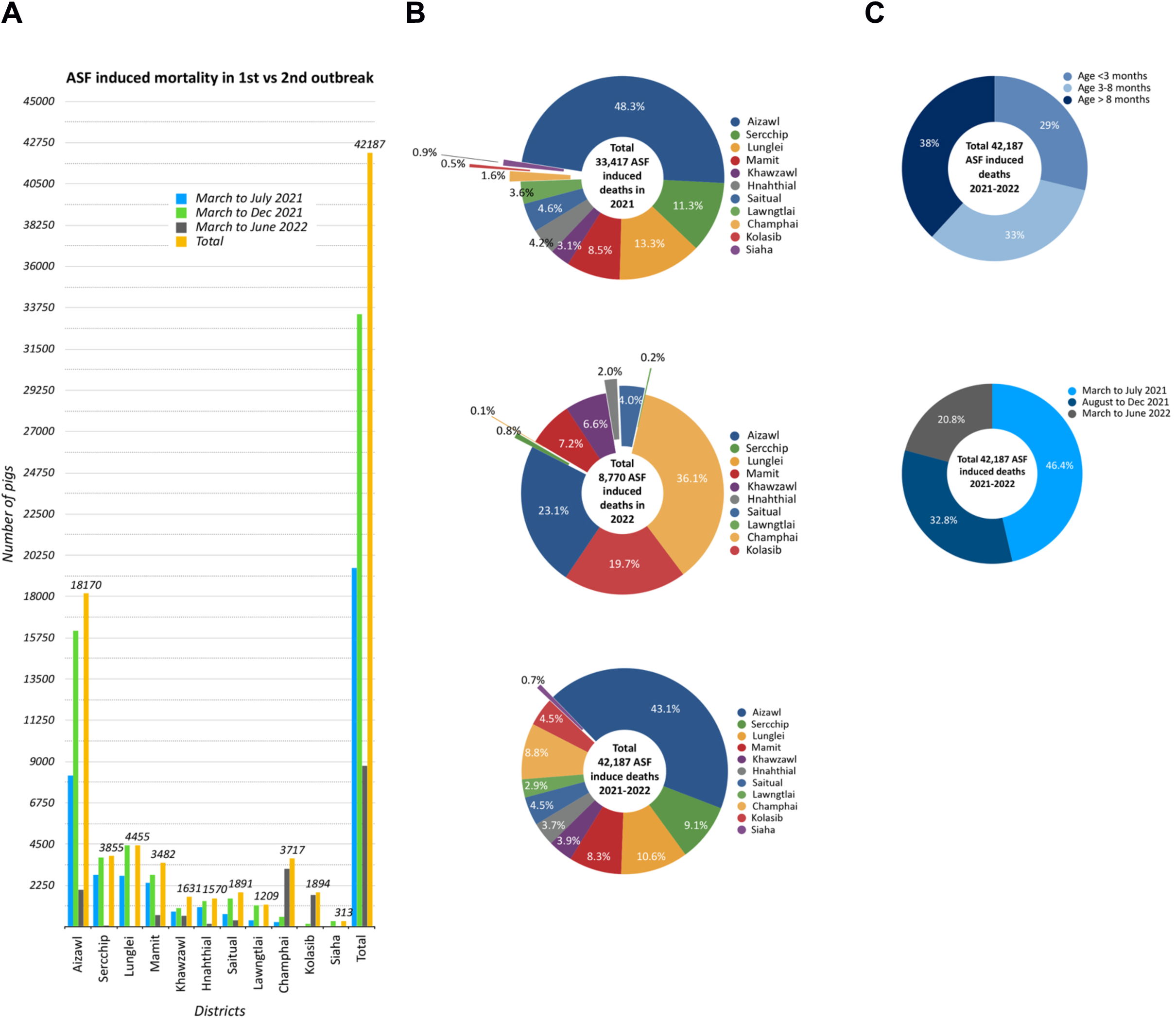
Overall ASF induced mortality in outbreaks between 2021 and 2022. A- District-wise mortality, B- Proportion of each district in overall mortality observed in Mizoram, C- Proportion of age group and outbreak period in the overall mortality.

A total of n=12,518 families were affected from the first and second ASF outbreaks of ASF, of which 77.3% (n=9,672) were affected in the first outbreak in 2021 while 22.7% (2,846) in the second outbreak in 2022. The district-wise proportion of families affected from the first and second outbreaks are presented in figure 5.

**Figure 5:**
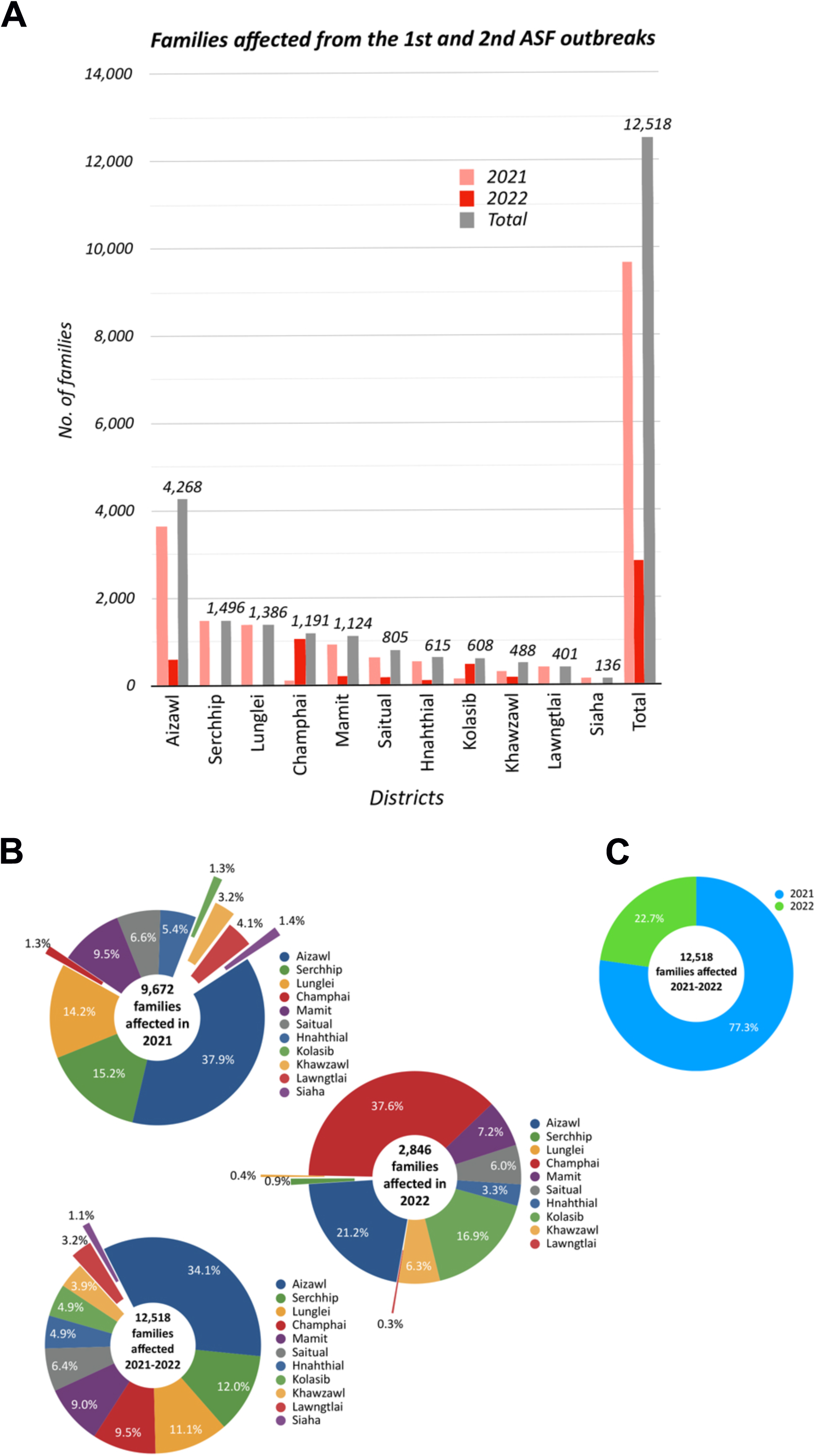
Livelihood affected from ASF outbreaks in Mizoram between 2021 and 2022. A- District-wise number of families affected, B- District-wise proportion of families affected, C- Year-wise contribution in total families affected.

## Discussion

Mizoram is one of major pig rearing Indian states contributing to the 47% of the total Indian pig population from the Northeast region [3,2,9]. Backyard or small holder pig farms constitute the major portion of piggery in the state while government and other commercial farm types are less [19, 15]. Therefore, swine diseases are undoubtedly a significant threat to the local livelihood in Mizoram, and the emergence of two fatal transboundary viral diseases - PRRS and recently ASF in less than 8 years gap not only destabilized the state’s economy but also induced psychosocial trauma [15, 20, 18]. In this article, we have attempted to dissect the account of ASF outbreaks that occurred for the first time in Mizoram using the data acquired from the Department of Animal Husbandry and Veterinary, Government of Mizoram. According to the data, we found that the first onset in the state was recorded on March 9, 2021, at Keitum village in Serchhip district. In the same month, ASF emerged at 1 location each in 5 other districts - Lunglei on the 21st, Lawngtlai on the 23rd, Siaha on the 26th, Mamit on the 28th and Aizawl on the 29th. Khawzawl and Kolasib reported their first onset in the month of April, Champhai in May and Hnahthial in June. By the end of July 2021 ASF had emerged in multiple locations in all 11 districts after showing a peak in the month of June, and the disease continued to spread till the end of 2021 before re-emergence in March 2022. Patil *et al* [21] reported similar observations on pattern of close ASF emergence within the first 3 months of ASF outbreak in Assam and Arunachal Pradesh and hypothesized such outbreaks as single cluster. They further speculated that the reason for rapid spread of ASF within this period to be “panic among the farmers thereby opting for distress sale of pigs at less rates”. We cannot rule out similar events contributing to the ASF peak in Mizoram even with the early advisory notice and efforts from the Government on compliance to biosecurity measures detailed in the National Action Plan on ASF [22, 23].

First laboratory confirmations by real-time PCR of the outbreaks in March 2021 in Serchhip, Lunglei and Aizawl were received on April 15, 2021, from the ICAR-National Institute of High Security Animal Diseases (NIHSAD), Bhopal, while laboratory confirmation for the ASF cases in Mamit was obtained from Northeast Regional Disease Diagnosis Laboratory, Guwahati. Confirmation of the ASF cases in Lawngtlai and the rest of the districts and following outbreaks were acquired from Department of Veterinary Microbiology, College of Veterinary Science and Animal Husbandry, Central Agricultural University, Aizawl. For the outbreaks in March 2021 the average duration from onset to specimen submission and specimen submission to laboratory confirmation were 13.2 days and 10 days respectively which is 23.2 days in average from onset to laboratory confirmation (data not shown). In an exceptional case, the laboratory confirmation of the March outbreak in Lawngtlai district was received after duration of 76 days despite the time taken for laboratory diagnosis by Department of Veterinary Microbiology was only 7 days, because the specimen was submitted after 68 days from the reported onset date of ASF in the district. Interestingly, the highest mortality rate by the end of 2021 was observed in Lawngtlai district going from 26.1% by mid-2021 to 81% by end of 2021 when the average mortality rate by end of 2021 was only 48.2%. According to the advisories to Indian states on ASF outbreak by Department of Animal Husbandry and Dairying, Government of India (DADH, GOI) [23] the ICAR-NIHSAD, a reference laboratory in Bhopal was designated for laboratory confirmation of ASF, however the subsequent laboratory diagnosis was executed by the Department of Veterinary Microbiology, Central Agriculture University in Aizawl because of logistic limitations in sending samples by the state to the reference laboratory owing to Covid-19 restrictions. It is important to note that the onset of ASF in these districts were detected during the Covid-19 pandemic when inter and intra-state movement restrictions and other social restrictions were not entirely relaxed. This situation, in addition to the difficult hilly terrain, was an impeding factor in the timely coordination of specimen collection, dispatch and eventually causing delay in laboratory confirmation and also compelling the state to rely on the closest laboratory for diagnosis. Although delay in laboratory confirmation might have some contribution in the cumulative loss due to ASF, the overall first-time ASF situation could have been far different and complicated without the covid-19 restrictions keeping in view that ASF virus is highly contagious and resilient that it is easily disseminated through fomites, meat scraps and handlers [7, 3].

Even though the CFR in the outbreaks under current study was reaching upto 100% the average mortality rate was observed to be 27.6% by mid-2021 and 48.2% by end of 2021, showing an average increase of 25.3%. Mamit district showed the second highest ASF mortality rate of 40% by mid-2021 next to 41.8% of Lunglei, but showed the second least percent change of 8.9% by end 2021 next to 8.5% of Khawzawl. Mamit district also showed the least CFR of 76.5% when the average CFR was observed to be 96.1%. Except Mamit, the CFR remained more or less consistent among the districts, but the polarized pattern of mortality rates among the districts by end of 2021 may have been a result of multifactorial attributes or determinants that influences the infection rate rather than just the virulence of the virus strain - varying time of deaths among pigs and size of pig population, time of culling before ASF induced deaths, distances between farms, biosecurity practices etc. Time of culling may have also influenced the CFR estimate. For example, the culling rate in Mamit district was 26.8% by mid-2021 and 88.5% by end of 2021 - the highest among the districts and the culling activity may have removed several infected pigs that would have otherwise succumbed to ASF induced death at the time of recording the number of deaths in the district. Hence, the high culling rate may explain the low CFR and low percent change of mortality rate estimated for Mamit. It was also observed that Lawngtlai district which had 0% culling rate coincidentally showed 100% CFR and highest percent change of mortality rate. On the contrary, Khawzawl district which had 0% culling rate showed the lowest percent change of mortality rate. These varying observations among districts are suggestive of other determinants of mortality rate coming into play, for instance, the population size of susceptible pigs in Khawzawl was approximately half the susceptible population in Mamit which may have resulted in a slightly different dynamic of virus transmission.

We observed that pigs above 8 months of age showed the highest ASF induced mortality in comparison to the other two age groups. Since we could not measure the age-wise mortality rate and infection rate, it is unclear if the above observation was due to actual difference in age-specific manifestation or simply a reflection of the age group dominant at the time of the outbreaks. Similarly, breed-wise response to ASF also remains to be investigated as Mizoram rears different breeds consisting of imported purebred and crossbred pigs as well as indigenous ‘Zovawk’, with 80% of the total population constituted by crossbred [15]. Nevertheless, ASF is of threat to conservation of germ plasm of ‘Zovawk’.

Information on the second outbreak in 2022 was limited to the cumulative age-wise deaths in each affected district, therefore, the mortality rate of 2022 could not be calculated. However, comparing the pig mortality during March to July 2021 and March to June 2022, the mortality in 2022 was 55.2% less than in 2021. This reduction in pig mortality in 2022 ASF outbreak was observed despite relative relaxation of Covid-19 restrictions and therefore may be attributed to increased biosecurity and other control measures in the second outbreak. However, district-wise comparison of mortality between outbreaks in 2021 and 2022 revealed that Champhai district contributed significantly (36.1%) in overall ASF-induced mortality in 2022 in contrast to 1.6% contribution to overall ASF-induced mortality in 2021. It is the highest of the only two districts showing spike in the second outbreak, the other being Kolasib contributing 19.7% in overall mortality in 2022 versus 0.5% in 2021. These findings can be correlated to the fact that Champhai district lies in the Indo-Myanmar border on the east, and while sharing a biodiverse buffer zone with Myanmar the district is also an informal trade corridor or ‘porous border’ for unregistered live pig and pork trade [15, 20]. Kolasib district is also a border district adjoining Cachar district and Hailakandi district of Assam in the north and northwest, and the main portal connecting Mizoram to the rest of India.

So far, two evidence hint the source of ASF in Mizoram and other states of India based on the genetic analysis of p72 and p54 encoding genes and central variable region of B602L gene. Rajkumar *et al.* showed the genetic similarity of Indian ASF virus associated with the outbreaks in Assam and Arunachal Pradesh to post-2007-p72-genotype II virus reported from Asia and Europe implicating that the Indian ASF was an event of transboundary expansion of the Asian ASF [10]. Rajkhowa *et al* characterized the ASF virus associated with the Mizoram outbreak also as genotype II. However, the information on B602L gene suggested that the virus was closely linked to the pre-2014 Eurasian strains and therefore, the Mizoram ASF virus and the other Indian strains belonged to two separate groups which, according to the authors, indicated that Mizoram ASF has a source different from the other states [18]. Nevertheless, the overall ASF morbidity rate and the mortality rate observed in Mizoram till July 2021 in our study were congruous with the observations of ASF till May 2020 in Assam and Arunachal Pradesh [21]. Although our study could not encompass the clinical profile of the disease during the outbreak, some of the clinical manifestations observed by Rajkhowa *et al* in the early outbreaks in Mizoram were severe depression, high fever, bloody diarrhoea, cutaneous haemorrhages and haemorrhagic lesions in visceral organs [18]. More structured and in-depth investigation on molecular epidemiology will not only reveal a clearer picture on the source of ASF virus but also explain the strain-driven basis of the clinical manifestations and mortality observed.

In a recent study in Assam, ASFV genotype-II was also detected in two carcasses of wild boar from two different national parks collected between 2020 and 2021 and the authors speculate the transmission of ASF virus from infected domestic pigs during the outbreaks in the region [24]. Moreover, there is an unofficial report stating the detection of 3 ASF cases in wild boar in Champhai district in July 2022 [25]. Although we are not entirely certain about the sources of these wild boar ASF cases, the infiltration of ASF virus among wild boar population irrefutably bears adverse consequences - not only as a threat to the wild life ecology but also as cradle for outbreak perpetuation, hence, further challenging the ASF mitigation policy as witnessed in countries in Africa, Europe and Asia [8, 12, 23]. With the existing poor implementation of biosecurity measures [21, 26] and unorganized live pig and pork trade through ‘porous border’ [15], the consequences of future threats are even more unfathomable in Mizoram.

We had also surveyed for ASF IgG in representative pig sera collected in the early 2022 from farms within the affected districts but did not observe any seropositivity against ASF (data not shown), which implies absence of live pigs with past exposure to ASF virus. However, this finding cannot truly represent the actual status due to the small sample size surveyed. The advisories to states on outbreak of ASF released in June 2019 by DADH, GOI when active outbreaks of ASF were reported in multiple Asian countries [23] and the formulation of National Action Plan on ASF in June 2020 [22] might have had some success on ASF management in the country. It is important to note that endemicity isn’t the only threat but the propensity of sporadic events leading to unprecedented outcomes is a factor that warrants revision of ASF control policies [12]. As unanimously pointed by other researchers [3, 21, 26], an extensive surveillance is pivotal toward achieving a more accurate assessments of risk factors and formulation of a practical and feasible mitigation plan tailored according to zones, and implementation thereof. Efforts to understand the ASF virus adaptation dynamic in the domestic host and wild boar in our region should also include studying the ecology of susceptible ticks and their possible roles. Continued concerted efforts between “Veterinary Services, wildlife and forestry authorities” as earlier emphasized by Sanchez et al [3] is of paramount need to manage the outbreaks in the country, and to mitigate the impact borne by the disease and risks imposed by ASF in wild boars.

Out of 62,895 susceptible pigs we estimated an overall 70% to have been depopulated by disease and culling by end of 2021. We do not have the information on the status of the remaining 30% pigs although we assume it to be live pigs. Whether these remaining pigs belonged to the same herd of affected pigs and likely acquired infection but were missed from the culling program and, if so, contributed to ASF re-emergence in 2022, or belonged to different unaffected herds or farms outside the culling radius is unknown to us. So far, in the absence of ASF vaccine and treatment, the most effective control measure appeared to be depopulation of susceptible pigs by culling [8]. It is understandable that 100% depopulation is not an economically viable option for ASF management in a state like Mizoram or other states in the country and, hence, as according to *Patil et al*, ASF is “a permanent threat to Indian pigs” [21].

So far, genotype-II has been dominant outside of African continent and a substantial progress on the efforts to develop effective vaccines against this genotype have been reported [27,28,29,30]. However, the tendency of other pig-adapted genotypes to expand in unfathomable geographical range demands a focus on broad spectrum vaccine with stringent biosecurity measures as previously highlighted in context to circulating ASF virus in African continent [28].

## Conclusion

We have presented an account on first ASF emergence in Mizoram, a small northeast state of India where pig farming is of major cultural and economic significance. The ASF impact is undoubtedly catastrophic and demands tailored and stringent biosecurity measures. Considering the nature and history of virus ecology, efforts on continued surveillance for risk factor analysis is of paramount importance.

## Data Availability

All data produced in the present work are contained in the manuscript

## Acknowledgments

The study was conducted as part of the project titled “Establishment of a Consortium for One Health to address Zoonotic and Transboundary Diseases in India, including the Northeast Region” approved by the Department of Biotechnology, Govt. of India with Order No: BT/PR39032/ADV/90/285/2020 dated 06/08/2021.

## Authors’ Contributions

SC conceptualised the study, curated and analysed the data, prepared the visualisation and wrote the original and final manuscript draft. AR, CN, EL and FL collected and compiled the data. PRC, PKS and TKD provided the resources. TKD supervised the study and reviewed the manuscript draft.

